# COVID-19 vaccines and autoimmune disorders: A scoping review protocol

**DOI:** 10.1101/2023.09.03.23295001

**Authors:** Claudia Chaufan, Laurie Manwell, Camila Heredia, Jennifer McDonald

## Abstract

**Background:** Two years into the global vaccination program, important questions about the association between COVID-19 vaccines and autoimmune diseases have arisen. A growing number of reports have documented associations between COVID-19 vaccination and autoimmunity, suggesting, for example, a causal link between vaccination and new-onset and/or relapsing autoimmune disorders such as type 1 diabetes mellitus, rheumatoid arthritis, multiple sclerosis, systemic lupus erythematosus, Graves disease, and Hashimoto’s thyroiditis. These autoimmune phenomena have occurred with various COVID-19 vaccines and research is required to elucidate the underlying mechanisms and causal directions, for example, whether persons with no history of autoimmune disorders may experience them upon vaccination or persons with autoimmune disorders may experience exacerbation or new adverse events post-vaccination.

**Methods and analysis:** Specific objectives of this scoping review will address the following questions: Can COVID-19 vaccination trigger and/or exacerbate autoimmune disorders? Are persons with autoimmune disorders at higher risk of experiencing additional autoimmune disorders? What are the mechanisms connecting autoimmune disorders with COVID-19 vaccination? Can COVID-19 vaccination interact with immunosuppressive therapy in persons with autoimmune disorders? Does the risk of autoimmune disorders following COVID-19 vaccination differ by vaccine type, age, gender, or other still unidentified characteristics (e.g., SES)? What is the consensus of care concerning COVID-19 vaccination in persons with autoimmune disorders and what evidence informs it? Our review will follow Arksey and O’Malley’s (2005) framework, enhanced by Levac et al.’s team-based approach (2010), and adhering to the recommendations of the Preferred Reporting Items for Systematic reviews and Meta-Analyses extension for Scoping Reviews (PRISMA-ScR) Checklist. To capture the broadest range of perspectives on the phenomenon of interest, data will be synthesized through numerical summaries describing general characteristics of included studies and thematic analysis. Subgroup analysis of primary outcomes will be performed to compare findings according to 1) the previous existence of autoimmune disorder, 2) the presence of relevant co-morbidities, 3) vaccine type; and other relevant factors that we may encounter as the research proceeds.

**Significance:** COVID-19 has triggered the largest vaccination campaign in history, targeting literally the global human community. Drug safety is a crucial aspect of any medical intervention, critical to a proper assessment of the balance of risks and benefits. Our investigation should yield information useful to improve medical and public health practice in multiple ways, including assisting in clinical decision-making, policy development, and ethical medical practice.

## 1. Background

### 1.1. The problem or issue

Since the launch of the global COVID-19 vaccination campaign in December 2020, there has been a slow but increasing recognition of adverse events post-inoculation. For instance, a July 2023 article in the peer-reviewed journal *Science* admitted to “apparent” complications, including “a debilitating suite of symptoms that resembles Long COVID” - among others, postural orthostatic tachycardia syndrome (POTS), abnormal blood clotting, heart inflammation, muscle weakness, fatigue, persistent headaches, and brain fog (1)(p. 18). While the list of what the authors refer to as “Long Vax” does not include autoimmune disorders, the literature reveals several areas of concern involving these disorders and COVID-19 inoculations. For example, already in 2021, Rocco et al. reported on the onset of autoimmune hepatitis upon COVID-19 vaccination in an elderly woman with a history of Hashimoto’s thyroiditis (2). That same year McShane et al. reported on a similar case, also in an elderly patient, this time with no prior history of autoimmune disorders (3). Two years into the global vaccination program, an observational, cross-sectional, pharmacovigilance cohort study examined individual case safety reports from VigiBase, the World Health Organization’s pharmacovigilance database, and identified a statistically significant association between COVID-19 vaccines and transverse myelitis, a condition that, as per the NIH, involves autoimmune processes (4). Finally, more recent case reports, including one of a healthy, 14-year-old girl deceased upon a third dose of an mRNA COVID-19 vaccine, described death as resulting from a fatal multiorgan inflammation activation, with multiple immune pathways potentially involved (5). Another case report described the onset of type 1 diabetes post second dose of the COVID-19 vaccine in a middle-aged woman with no prior history of the disease (6).

There is also a dearth of ongoing research on persons experiencing autoimmune disorders. Indeed, public health agencies and medical organizations strongly endorse COVID-19 vaccination among these persons and assert that the benefits outweigh what are considered rare risks (7–10). However, a search of scoping reviews in the Cochrane Collaboration and Joanna Briggs websites, and of systematic reviews in the PROSPERO website, conducted on August 22, 2023, revealed zero and 7 planned studies, respectively, examining the relationship between COVID-19 inoculations and autoimmune disorders, none of them focusing on the six most frequent disorders: type 1 diabetes, rheumatoid arthritis, lupus, multiple sclerosis, Hashimoto’s thyroiditis, and Graves’ disease (11) and autoimmune disorders more generally. As well, the mechanisms linking inoculations and these disorders - whether the former increases the risks of autoimmune adverse events, or whether patients with autoimmune disorders experience exacerbation or new adverse events post-vaccination - are poorly understood and documented, thus our planned review.

### 1.2. Description of the phenomenon of interest

Systematic or scoping reviews generally evaluate “interventions”. However, these approaches can be applied productively to reviews that seek to document and critically appraise broader phenomena, i.e., “phenomena of interest”, from diverse sources. Our phenomenon of interest is the evidence for bidirectional relationships and underlying mechanisms between COVID-19 inoculations and autoimmune disorders.

### 1.3. Why it is important to do this review

The relationship between vaccination in general and autoimmunity is well documented, and many potential mechanisms have been hypothesized and demonstrated, in the lab and in the clinic. Autoimmunity is characterized by abnormal immune system responses directed against the host rather than a foreign invader and can lead to the production of autoantibodies and the development of autoimmune diseases such as type 1 diabetes mellitus (T1DM), rheumatoid arthritis (RA), multiple sclerosis (MS), systemic lupus erythematosus (SLE), Graves disease, and Hashimoto’s thyroiditis (12). Associations between vaccination and autoimmunity have been identified for the influenza vaccine and Guillan-Barré syndrome, the oral polio vaccine and transverse myelitis, and combination vaccines such as diphtheria-tetanus-pertussis (DTP) and measles-mumps-rubella (MMR) and arthritis (13). Similarly, review reports suggest a causal link between COVID-19 vaccination and new-onset and/or relapsing autoimmune disorders including SLE, RA, T1DM, Hashimoto’s thyroiditis, and Graves’ disease (14–16). For example, “new-onset cases have been reported within weeks of COVID-19 vaccination, 12 cases of new-onset SLE were documented, 7 cases of new-onset RA were reported characterized by morning stiffness, swelling, pain, and positive rheumatoid factor (RF) serological tests, and 13 cases of new-onset T1DM were reported with symptoms including excessive thirst, urination, and fatigue, and symptom reversal with insulin treatment (14). These autoimmune phenomena have occurred with various COVID-19 vaccines including adenovirus vector and novel mRNA vaccines (17,18).

As noted in the *Science* article mentioned earlier, the “Post-COVID” or “Long COVID” and the “Long Vax” syndromes share a range of symptoms, such as persistent cough, shortness of breath, and chest pain, severe and chronic fatigue, sleep disorders, headaches, and cognitive impairments, including compromised concentration and memory loss (18,19). In both cases, these symptoms could be related to the accumulation of SARS-CoV-2 spike protein in blood vessel epithelium compromising the blood-brain-barrier (BBB) and leading to signs of encephalomyelitis/chronic fatigue syndrome (ME/CFS) (19). It follows that COVID-19 inoculations could operate through spike protein production that further compromises neuroimmunoendocrine function by producing novel autoantibodies (18).

#### Mechanisms linking COVID-19 inoculations and autoimmune disorders

It has been proposed that viral infections cause autoimmunity and thus it is likely that similar mechanisms are involved in the link between vaccinations and autoimmunity (13). Mechanisms of autoimmunity can be antigen-specific, non-specific, or a combination of both. The most commonly occurring autoimmune events post-vaccination include autoantibodies, encephalitis, neuropathy, demyelination, vasculitis, and arthritis (13). Potential mechanisms for the relationship between vaccination and autoimmunity include 1) molecular mimicry; 2) bystander activation; 3) epitope spreading; 4) polyclonal activation of B cells; 5) adjuvant-induced immune stimulation, and other adverse events that aggravate immune system responses (13,14). *Molecular mimicry*, an antigen-specific mechanism, occurs when foreign antigens are recognized as host antigens, such as when lipopolysaccharides and peptides produced by microorganisms mimic those of the host in the immune, nervous, and endocrine systems (12).

*Bystander activation*, an antigen-*non*-specific mechanism, occurs whenever infection triggers innate immune response or the release of self-antigens leading to activation of autoreactive T cells, cytokine secretion, inflammation, and more autoreactive lymphocytes (12). *Epitope spreading (ES)* occurs when the immune response to an epitope differs from the dominant epitope and lacks cross-reactivity. Therefore, the immune response can then spread by intramolecular diffusion (i.e., to other epitopes on the same protein) or by intermolecular diffusion (i.e., to epitopes on different proteins) (14) *Polyclonal activation* occurs when there is chronic immune system activation leading to the proliferation of B cells and immune complex formation circulation (14).

Finally, *vaccine adjuvants*, i.e., compounds added to vaccines as delivery systems or immune potentiators, have also been shown to cause adverse reactions, referred to collectively as *autoimmune/inflammatory syndrome induced by adjuvants* (ASIA) (20). Of note, dozens of patients have been diagnosed with ASIA post-COVID-19 vaccination (19). The mechanism of action of adjuvants in ASIA varied and include enhanced uptake of antigens, inflammasome activation, increased cell recruitment at the site of injection, and toll-like receptor activation (20). Various adjuvants linked to ASIA include aluminum-based materials, mineral oils, and silicone compounds (20,21).

#### Autoimmunity, the Neuroimmunoendocrine System, and COVID-19 inoculations

Yet another understudied area is the relationship between COVID-19 inoculations and the neuroimmunoendocrine system. Psychoneuroimmunoendocrinology (PNIE) focuses on the relationship between structural/functional (nervous, immune, and endocrine systems) and psychological (e.g., cognition, emotion, and behaviour) aspects of the human organism and is important to address the etiology and treatment of immune-mediated diseases for two reasons: 1) many immune-mediated diseases are characterized by significant dysregulation in multiple organ systems, including high expression of proinflammatory cytokines, alterations of the blood-brain barrier (BBB, neuroinflammation, and changes in anabolic and/or catabolic hormones; 2) many immune-mediated diseases involve feedback mechanisms wherein psychological stressors can induce and/or exacerbate autoimmune disorders that in turn create significant psychological stress (22). Thus, PNIE is critical to understand autoimmunity during the COVID-19 crisis because of the significant and widespread psychological stressors, such as social isolation, experienced by populations following public-health countermeasures like prolonged quarantines and mass lockdowns (23–26).

SARS-CoV-2 infections crossing the BBB and invading neurons and glia expressing angiotensin-converting enzyme 2 (ACE2) in the hypothalamic-pituitary region may cause autoimmune responses that can precipitate and/or exacerbate the hypothalamic-pituitary-adrenal (HPA) stress response system (22). Corticoids released in the stress response also play an important role in preventing damage to tissues through a negative feedback system that works to suppress the production and circulation of cytokines, including tumor necrosis factor α (TNF-α), interleukin 1 beta (IL-1β), and interleukin 6 (IL-6) involved in COVID-19 pathogenesis (22,27,28). Specifically, it is thought that the virus disrupts normal HPA function through the induction of anti-adrenocorticotropic hormone (ACTH) antibodies leading to reduced efficacy of the body’s immune response (22).

The proposed mechanism for HPA autoimmunity is a molecular mimicry of amino acid sequences of the host ACTH (22). SARS-CoV-2 can also induce mast cell activation which has been associated with neuroinflammation, neurodegeneration, and psychological stress (28). As well, increased neuroinflammatory responses could exacerbate pathology in individuals affected by other immune-mediated diseases such as multiple sclerosis (MS), amyotrophic lateral sclerosis (ALS), Huntington’s disease, Alzheimer’s disease (AD), and Parkinson’s disease (PD) (28,29). Dysregulation of the neuroimmunoendocrine system, which is critical in maintaining homeostasis during periods of stress, helps to explain the greater vulnerability of individuals with immune-mediated diseases and immunosenescence to the combination of SARS-CoV-2 infection and significant psychological distress arising from governmental responses to the pandemic.

In concluding, COVID-19 has triggered the largest vaccination campaign in world history, targeting literally the global human community. Drug safety is a crucial aspect of any medical intervention, critical to a proper assessment of the balance of risks and benefits. Our investigation should yield information useful to improve medical and public health practice in multiple ways, including assisting in clinical decision-making, policy development, and ethical medical practice.

## 2. Objectives

Our broad objective is to appraise what the existing literature reports about the association between COVID-19 vaccines and autoimmune disorders, in general, and specifically concerning type 1 diabetes (T1DM), multiple sclerosis (MS), Hashimoto’s thyroiditis, rheumatoid arthritis (RA), lupus, and Graves’ disease. Specific objectives will address the following questions: 1) In persons with autoimmune disorders, general or specific – type 1 diabetes mellitus (T1DM), rheumatoid arthritis (RA), multiple sclerosis (MS), systemic lupus erythematosus (SLE), Graves disease, and Hashimoto’s thyroiditis - can COVID-19 vaccination *trigger* autoimmune disorders? 2) In persons with autoimmune disorders, general or specific – type 1 diabetes mellitus (T1DM), rheumatoid arthritis (RA), multiple sclerosis (MS), systemic lupus erythematosus (SLE), Graves disease, and Hashimoto’s thyroiditis - can COVID-19 vaccinations *exacerbate* autoimmune disorders? 3) Are persons with autoimmune disorders at higher risk than persons without autoimmune disorders of experiencing additional autoimmune disorders? 4) What are the mechanisms connecting autoimmune disorders with COVID-19 vaccination? 5) Can COVID-19 vaccination interact, positively or negatively, with immunosuppressive therapy in persons with general or specific autoimmune disorders under treatment with this therapy? 6) Does the risk of general or specific autoimmune disorders following COVID-19 vaccination differ by vaccine type, age, gender, or other still unidentified characteristics (e.g., SES)? and 7) What is the consensus of care concerning COVID-19 vaccination in persons with general or specific autoimmune disorders what evidence informs it?

## 3. Methods

### 3.1. General considerations

To address our review objectives and achieve our research objective, we will conduct a scoping review of the literature following Arksey and O’Malley’s framework (30). These authors propose that, in contrast to systematic reviews that “typically focus on a well-defined question where appropriate study designs can be identified in advance [and] provide answers to questions from a relatively narrow range of quality assessed studies”, scoping reviews help to “address broader topics where many different study designs might be applicable [and are] “less likely to seek to address very specific research questions nor, consequently, to assess the quality of included studies” (p. 20), thus our choice of this review approach. Our analysis will be enhanced by Levac et al.’s team-based approach (31) that proposes that throughout the review, from articulating a research question, identifying, and selecting relevant studies, charting the data and collating, summarizing, and reporting results the process should be iterative and cooperative, i.e., “team-based”. This approach should help to address unforeseen practical challenges, such as the need to refine inclusion/exclusion criteria as during the screening and selection process.

### 3.2. Protocol registration and reporting

This protocol was registered with the Open Science Framework (blinded). The scoping review will follow the recommendations of the Preferred Reporting Items for Systematic reviews and Meta-Analyses extension for Scoping Reviews (PRISMA-ScR). Upon registration of the protocol, an eight-month timeframe will be dedicated to the search, selection, data extraction, and writing of the scoping review. This will result in a tentative completion date of the manuscript by April 2023.

### 3.3. Ethics review

Because the data are publicly available, no IRB approval is required.

### 3.4. Criteria for considering studies for this review

#### 3.2.1 Types of studies

This scoping review will include articles in English that refer to the association between any type of COVID-19 vaccine and autoimmune disorders (listed under objectives) reporting on empirically verifiable clinical manifestations of autoimmunity, with no temporal or geographic restrictions, in populations of any age, sex/gender/race/ethnicity, socioeconomic class, or national origin, accessible through the libraries of the authors’ academic / professional affiliations.

#### 3.2.2 Types of interventions

To capture the broadest range of perspectives on the association between COVID-19 vaccines and autoimmune disorders, all articles, regardless of type (e.g., original article, case report, technical note) will be included, provided they meet the inclusion criteria (32).

#### 3.2.3 Types of outcome measures

Outcomes will be tailored to capture the phenomenon of interest. They will include side effects, relapses/flares of previous autoimmune diseases, and new clinical autoimmune or other manifestation of the association between any COVID-19 vaccine and autoimmune disorders.

### 3.5. Search methods for identification of studies

#### 3.3.1 Electronic searches

We will retrieve data from 1) PubMed, and 2) WHO (World Health Organization) database. For PubMed, we will use the search MeSH Major Topic terms [Hashimoto’s Thyroiditis” OR “Graves disease” OR “rheumatoid arthritis” OR “type 1 diabetes” OR “systemic lupus erythematosus” OR “multiple sclerosis” OR “autoimmunity” OR “autoimmune diseases” OR ‘‘autoimmune disorders’’]. These terms will be combined with [“COVID-19 vaccines”]. For the WHO search, the same terms will be searched as [Title, abstract, subject]. At the outset of the search, specific eligibility criteria will include the following: i) between 2000 and 2023, ii) English only, and ii) pre-print or published.”

#### 3.2.2 Searching other sources

Reports by leading national and international health agencies – e.g., Centers for Disease Control and Prevention, and World Health Organization – will be used to contextualize the study.

### 3.6. Data collection and analysis

#### 3.4.1 Description of methods of selected-in studies

To capture the broadest range of perspectives on the phenomenon of interest, this scoping review will include all articles, regardless of type or methodology (e.g., original research, case report, technical note), that report on the association between any COVID-19 vaccine and autoimmune disorders (as listed under “objectives”) and meet the inclusion criteria.

#### 3.4.2 Study selection and inclusion criteria

Before including articles for assessment, we will conduct a preliminary screening of the literature search to discard irrelevant material. One reviewer will initially scan titles and remove those who do not meet the inclusion criteria: 1) articles on the possible or actual association between any type of OVID-19 vaccine and any type of autoimmune disorder 2) focus on empirically verifiable clinical manifestation of autoimmune disorders 3) available through any of the library websites of author’s affiliated universities; 4) in English. Next, two reviewers will independently scrutinize the remaining abstracts in relation to review objectives (as described earlier) and eliminate those that meet any of the exclusion criteria: 1) not on COVID-19 2) not on vaccines 3) not on autoimmunity 4) no identifiable clinical outcome 5) no original or primary data 6) not in English.

Where there is uncertainty in the abstract about the relevance of an article, a third reviewer will break the tie, and if needed, the full text will be retrieved. Once the abstract review process is complete, we will retrieve full copies of the selected articles for assessment. Two reviewers will independently determine if the articles meet the inclusion criteria and will screen them independently. Disagreements will be resolved by full team discussion.

We will monitor inter-rater reliability throughout the screening stage on a regular basis (after about one-fourth of retrieved articles are screened), and act if the reliability falls below 80% (33). We will report these scores in the final review, maintain a clear record of the articles included and excluded at each stage of the process, and note the reasons for excluding specific articles. Articles that do not meet the inclusion criteria but include relevant contextual material (e.g., policy papers) may be narratively summarized in the background section of the final manuscript.

Throughout the screening process, we will use the Rayyan literature review management software (https://www.rayyan.ai/) to 1) facilitate double-blind screening, 2) record inclusion and exclusion decisions, and 3) identify any disagreements between reviewers.

#### 3.4.3 Data charting

The data charting form will be prepared using Microsoft Excel. Chartered data will include details about article type; data informing our phenomenon of interest (e.g., autoimmune manifestation; vaccine type; age of patient; clinical background); and contextual factors (e.g., country where event was reported). Data charting will be performed by two researchers. Before beginning full charting, two reviewers will independently chart data from a common sample of studies and the team will meet to calibrate the approach and discuss results. Tables for all included articles will be created and included as appendices in the final review.

#### 3.4.4 Assessment of risk of bias in included studies (we will not do this but need to explain why)

Scoping reviews chart the evidence concerning a particular subject, pinpoint core concepts, theories, sources, and areas where gaps in knowledge exist, and offer a comprehensive outlook on existing evidence, without considering the methodological quality of charted articles. As a result, sources of evidence in this scoping review will not be subjected to quality assessment.

#### 3.4.5 Data synthesis

In convergent synthesis designs, data is transformed into either qualitative or quantitative findings. In convergent *qualitative* synthesis, our chosen approach, results from diverse methodologies are transformed into qualitative findings such as concepts, patterns, and themes (34,35). In this scoping review, data will be synthesized through descriptive numerical summaries, describing general characteristics of included studies, and thematic analysis (30,36).

#### 3.4.6 Subgroup analysis

Subgroup analysis of primary outcomes will be performed to compare findings according to 1) the previous existence of autoimmune disorder; 2) the presence of relevant co-morbidities (37); 3) vaccine type; and other relevant factors that we may encounter as the research proceeds.

## Data Availability

This is a review protocol, with no original data

https://osf.io/n4gxz

## Author contributions

CCh designed the scoping review, wrote the protocol, and will oversee, and participate in, every step of the project until the completion and publication of the umbrella review. LM assisted with the study design and protocol drafting, provided expertise in neuroimmunoendocrinology, and will participate in article selection, data charting / synthesis / analysis, and final review writing. CH assisted with the study design, provided expertise in methodology, conducted the first round of data screening, and will participate in article selection, data charting / synthesis / analysis, and final review writing. JM assisted with the study design and will participate in article selection, data charting / synthesis / analysis, and final review writing. All the authors have read and approved the final version of this protocol.

## Source of support

This work has been partially funded by a New Frontiers in Research Fund (NFRF) 2022 Special Call, NFRFR-2022-00305.

## Declarations

Ethics approval and consent to participate – not applicable. Consent for publication – not applicable.

## Competing interests

The authors have no conflicts of interest to declare. Funders and professional / academic affiliations have played no role in the conception, conduct, or decision to conduct this research or submit it for publication.

## Notes

### Competing Interest Statement

The authors have declared no competing interest.

